# Intrathecally expanded GZMK+/GZMH+ CD8 T cells targeting EBV antigens may reduce severity of Multiple Sclerosis

**DOI:** 10.1101/2025.08.05.25333071

**Authors:** Shinji Ashida, Peter Kosa, Francisco Otaizo-Carrasquero, Dan Sturdevant, Amirhossein Shamsaddini, Yongmei Zhao, Jyoti Shetty, Craig Martens, Jeffrey I. Cohen, Bibiana Bielekova

**Affiliations:** Neuroimmunological Diseases Section, Laboratory of Clinical Immunology and Microbiology, National Institute of Allergy and Infectious Diseases, National Institutes of Health, Bethesda, MD, USA; Genomic Research Section, Research Technologies Branch, National Institute of Allergy and Infectious Diseases, National Institutes of Health, Bethesda, MD, USA; Sequencing Facility Bioinformatics Group, Advanced Biomedical and Computational Science, Frederick National Laboratory for Cancer Research, Frederick, MD, USA; Medical Virology Section, Laboratory of Infectious Diseases, National Institute of Allergy and Infectious Diseases, National Institutes of Health, Bethesda, MD, USA

## Abstract

Combining cerebrospinal fluid B cell receptor and T cell receptor repertoire analysis with transcriptional/ flow cytometry cellular profiles in hundreds of deeply-phenotyped people with Multiple Sclerosis (pwMS) and controls, we identified intrathecal expansion of anti-viral, cytotoxic, granzymes H/K (GZMH+/GZMK+) double positive (DP) CD8+ T cells that recognize EBV epitopes in pwMS. DP CD8+ T cells are activated and expanded by, and kill autologous, EBV-infected CSF B cell lines in-vitro. Correlations of surrogate transcriptional profiles with clinical and imaging outcomes infer a beneficial role for EBV-targeting DP CD8+ T cells, as untreated pwMS with proportionally higher DP CD8+ T cells to intrathecal B cells accumulate neurological disability slower. MS therapies also increase ratios of beneficial CD8+ T cell responses to intrathecal B cells, consistent with their ability to inhibit disability progression. This study provides indirect evidence that intrathecal EBV infection participates in disability accumulation in pwMS.

## Introduction

Multiple sclerosis (MS) is chronic immune-mediated disease of the central nervous system (CNS), characterized by formation of focal demyelinating lesions with accompanying neuro-axonal loss. While varied immunomodulatory treatments inhibit MS lesions formation, which immune cells damage CNS tissue, and how and why this occurs remains unanswered.

Partial resemblance to experimental autoimmune encephalomyelitis implicated myelin specific CD4+ T cells as contributing to MS. Their pathogenic role in pwMS is supported by genetic linkage to MHC-II^1,2^ and the observation that when altered peptide ligand therapy inadvertently expanded myelin-specific CD4+ T cells > 1000-fold, these cells caused focal CNS lesions^3^. But is that how MS lesions form naturally?

Initially described differences in myelin-specific T cells or antibodies (Ab) frequencies between pwMS and controls^4–6^ were not reproduced when controls included other inflammatory neurological diseases (OIND) or were MHC-II-matched to pwMS^7–10^. While new reports describing autoimmune responses in pwMS continue to emerge^11–13^, they have not been convincingly linked to clinical outcomes.

Virus-associated demyelination is an alternative and not mutually exclusive^13^ hypothesis supported by animal observations^14–16^. Indeed, epidemiology links Epstein Barr Virus (EBV) infection to MS risk^17–19^ and some^20–22^, but not all^23,24^ MS pathology studies identified EBV in MS CNS. As a ubiquitous herpesvirus, EBV establishes life-long latency in B cells where it episodically reactivates, especially upon differentiation to plasma cells (PC)^25^. EBV infects also epithelial cells^26^, causing lytic infection. Reproducible identification of intrathecal EBV infection may be difficult when intrathecal B cells are rare (<100/analyzed sample), their latent infection several logs rarer (between 1-10/100,000^27^), and lytic reactivation is unpredictably episodic and likely rapidly cleared by immunocompetent hosts.

In the absence of being able to quantify intrathecal EBV infection directly, detection of EBV infection requires indirect measurement, by quantifying anti-EBV host responses. Many studies reported increased Ab responses against EBV proteins^17^ in pwMS (reviewed^9^). Furthermore, increased CD8+ T cell responses against EBV antigens or EBV-transfected CSF B cells^28^ were observed in pwMS blood^29^, CSF^28,30,31^ or brain^32^.

While this collectively links EBV infection to MS, essential questions remain unanswered: 1. Does EBV only trigger MS, or does it participate in MS progression? 2. Do anti-EBV T cell responses contribute to CNS damage via molecular mimicry or bystander damage, or do they limit tissue injury by restricting intrathecal EBV reactivation? 3. If anti-EBV immunity plays a protective role in MS, what is its phenotype? This paper aimed to answer these questions.

## Results

### B cell receptor (BCR) and T cell receptor (TCR) repertoire analysis demonstrates intrathecal clonal expansion of B cells and T cells in pwMS

BCR and TCR libraries were generated in blinded fashion from 789 bulk RNA sequence reactions of cerebrospinal fluid (CSF) cells prospectively collected over 12 years from subjects presenting for diagnostic work-up or management of possible neuroimmunological disorder (Extended Data Table 1). 526 CSF samples (348 from pwMS, 14 from clinically isolated syndrome [CIS; subjects with initial clinical attack suggestive of MS but not yet fulfilling MS diagnostic criteria] and 164 from controls) contained sufficient BCR data (Figure 1A).

**Figure 1:**
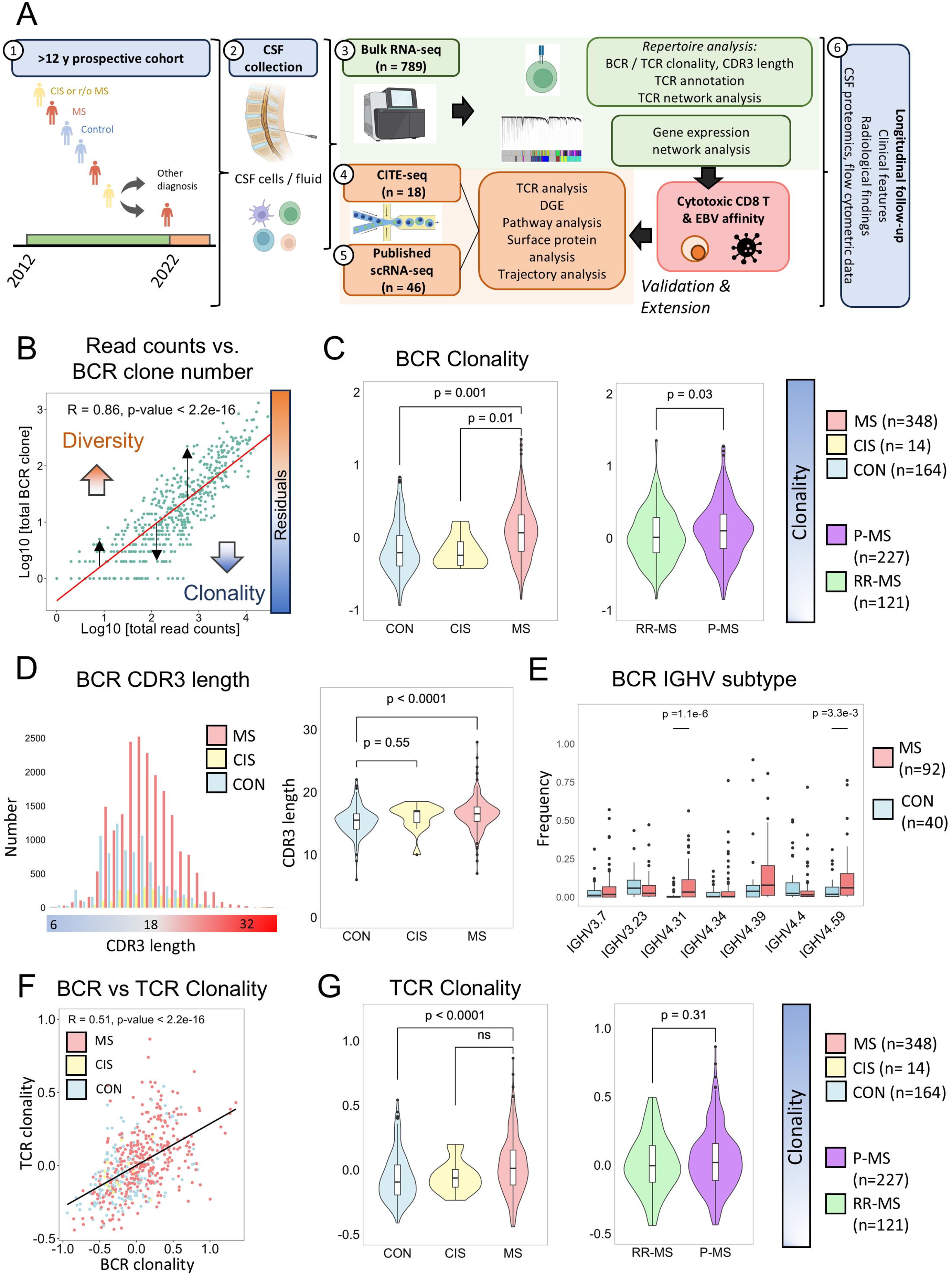
TCR and BCR repertoire analysis. A. Schematic representation of bulk RNA and single-cell RNA sequencing analysis of CSF cells B. Linear regression model depicting the relationship between total BCR read counts and total BCR clone numbers. Black arrows indicate the residuals of the model, where positive residuals suggest increased diversity, and negative one suggest clonality. The R value of the linear model and the p-values are shown in the plot C. Comparison of BCR clonality across MS, CIS, and control (left) and RR-MS and progressive MS (right) D. Distribution of BCR CDR3 length was plotted based on MS, CIS, and control (left) and compared among groups (right) E. Comparison of the dominant BCR IGHV subtypes between MS and control groups (of which BCR clone number ≥50), with IGHV4.31 and IGHV4.59 being more dominant in MS F. TCR clonality showed moderate correlation with BCR clonality G. Comparison of TCR clonality across MS, CIS, and control (left) and RR-MS and progressive MS (right). Mann-Whitney U test was used for comparisons between two groups, while Tukey-Kramer test was applied for comparisons among three groups. R indicated clustering coefficient.

We calculated BCR (and TCR) clonality by adjusting for total read counts (Figure 1B). After unblinding diagnostic categories, pwMS had significantly higher BCR clonality than controls (Figure 1C). BCR clonality was also higher in pwMS compared to CIS, and in people with progressive MS (P-MS), which represents a later MS stage in comparison to early, relapsing-remitting MS (RR-MS; Figure 1C). Thus, BCR clonality increases with MS progression.

PwMS had also significantly longer BCR complementarity determining region 3 (CDR3) compared to controls (i.e., median of 16.5 vs 15.5; p<0.0001, Figure 1D). Longer CDR3 in clonally expanded B cells are observed in both viral infections^33^ and autoimmunity^34^.

To infer whether intrathecal B cells target shared antigens, we assessed immunoglobulin heavy chain gene (IGHV) subclasses. We first identified the most expanded IGHV subtypes. A subsequent validation cohort (92 pwMS and 40 control CSF with ≥50 BCR clones) verified significantly higher IGHV4.31 and IGHV4.59 frequencies in pwMS compared to controls (Figure 1E).

These IGHV subclasses may be expanded in pwMS because they recognize shared antigen(s) or because they receive preferential help from CD4+ T cells restricted by MS-enriched DRB1*1501 (and DRB1*1503) alleles. Indeed, IGHV4.31 and IGHV4.59 frequencies were enriched in pwMS with DRB1*1501 or DRB1*1503 MHC-II alleles, although due to small sample sizes the difference did not reach statistical significance (Extended Data Figure 1A).

Analogous analyses of TCR clonality (Extended Data Figure 1B) found a correlation between TCR and BCR clonality (Figure 1F) and TCR clonality significantly increased in pwMS without differences between P-MS and RR-MS (Figure 1G).

We conclude that compared to controls, pwMS have intrathecal clonal expansion of B cells and T cells that is sustained (for TCRs) or further enhanced (for BCRs) during MS progression.

### TCR from MS CSF shows higher similarity and increased recognition of EBV lytic epitopes

To study if the frequency of intrathecal T cells recognizing EBV antigens differs between pwMS and controls, we used reference databases to identify CDR3 sequences known to recognize epitopes from 3 human viruses: cytomegalovirus (CMV), EBV and Influenza. Additionally, recognizing that TCR databases catalogue only a tiny fraction of anti-viral TCRs, we used CDR3 clustering to assess whether non-annotated TCRs have structural similarity (and thus likely bind to the same or similar antigens^35^) to TCRs annotated to specific viruses (Figure 2A).

**Figure 2:**
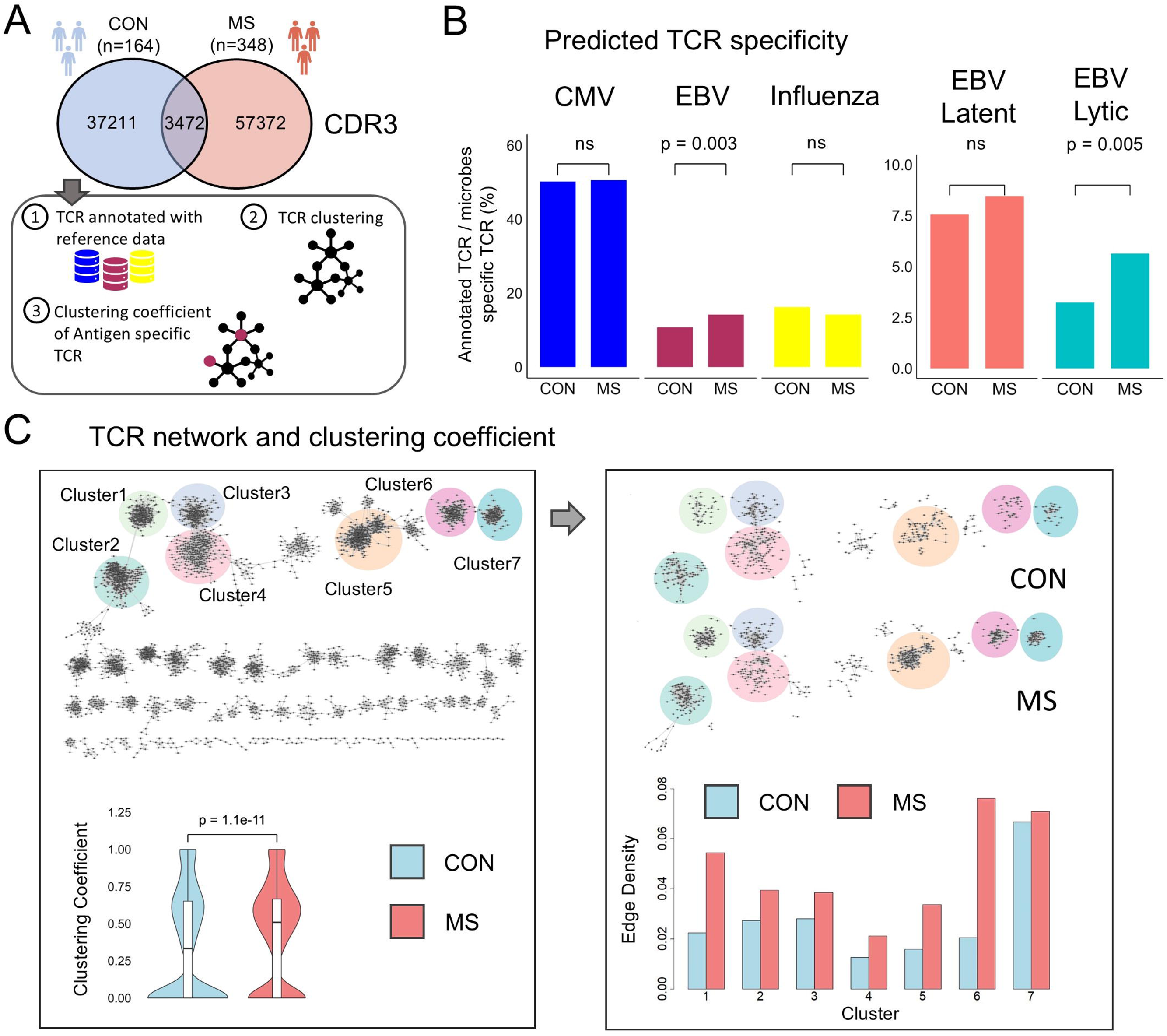
TCR similarity and prediction of TCR antigen specificity. A. The CDR3 library was generated from MS and control. CDR3 was annotated to specific antigen by using three CDR3 cohorts (VDJdb, McPAS-TCR, and TBAdb). MS, control specific and shared CDR3 library (each 1500 CDR3s) were randomly selected to generate CDR3 network. Then, TCRs annotated to each virus were superimposed with TCR clustering coefficient. B. The antigen annotation: The frequency of TCRs specific for CMV, EBV, and influenza were compared between MS and control. TCR from pwMS showed higher frequency of TCRs annotation to EBV (left). The frequency of TCRs annotation to lytic and latent EBV protein were assessed and compared between MS and control (right). Lytic protein was predicted to annotated higher in MS than control. C. Network similarity was assessed by clustering coefficient. CDR3 network in MS showed higher clustering coefficient (i.e., similarity) compared with control (left). In network analysis, the edge density of MS was higher, especially in clusters 1, 5, and 6 compared with control.

Of the CDR3 library, 2,671/57,372 (4.7%) in pwMS and 1,590/37,211 (4.3%) in controls matched published databases, without significant differences between cohorts (Extended Data Figure 1C). Likewise, cohorts did not differ in proportions of CDR3s recognizing CMV or influenza epitopes. In contrast, a significantly higher proportion of intrathecal TCRs overlapped with EBV-annotated CDR3 in pwMS (i.e., 379/2,671 =14.1% vs. 171/1,590 =10.8%, p = 0.003; Figure 2B, left). This was preferentially driven by increased TCR reactivity to EBV lytic epitopes (p=0.005, Figure 2B, right) in pwMS. Comparable findings were obtained when the analysis was restricted to HLA-A02-restricted TCRs (Extended Data Figure 2A).

Clustering algorithm based on the minimal number of single-character edits between CDR3s (see Methods) clustered all 4,500 CDR3s into 7 large and close to 50 small clusters (Figure 2C). PwMS had a significantly higher TCR clustering coefficient compared to controls (p=1.1e-11) and dominated some of the large TCR clusters (e.g., clusters 1, 5 and 6; Figure 2C, right).

Clustering coefficients for virus-targeting TCRs were comparable between pwMS and controls except a nonsignificant trend for a higher clustering coefficient of EBV TCRs in pwMS (Extended Data Figure 2B). We conclude that a higher TCR similarity in pwMS suggests partially overlapping targets, some of which may be EBV epitopes.

### TCR Similarity is Linked to Cytotoxic CD8+ T Cells in MS

Next, we combined the CSF cells bulk RNA-seq with flow cytometry-based enumeration of immune cell subtypes to search for T cells expanded in pwMS that target shared antigens based on TCR similarity.

First, we identified genes correlating with the TCR clustering coefficient. Due to large number of transcripts per samples, we randomly split (1:1) MS samples into Discovery and Validation cohorts (Figure 3A). Genes with a Spearman correlation coefficient ≥ 0.1 or ≤ −0.1 and p-value <0.05 (3627 genes) in the discovery cohort were analyzed in the validation cohort. 26 positively and 44 negatively correlated genes with TCR similarity achieved an FDR adjusted p-value < 0.05 (Figure 3B). As *CD8A, NKG7, GZMH* and *F2R* achieved the highest positive correlations, we conclude that cytotoxic CD8+ T cells have highest TCR similarity in pwMS.

**Figure 3:**
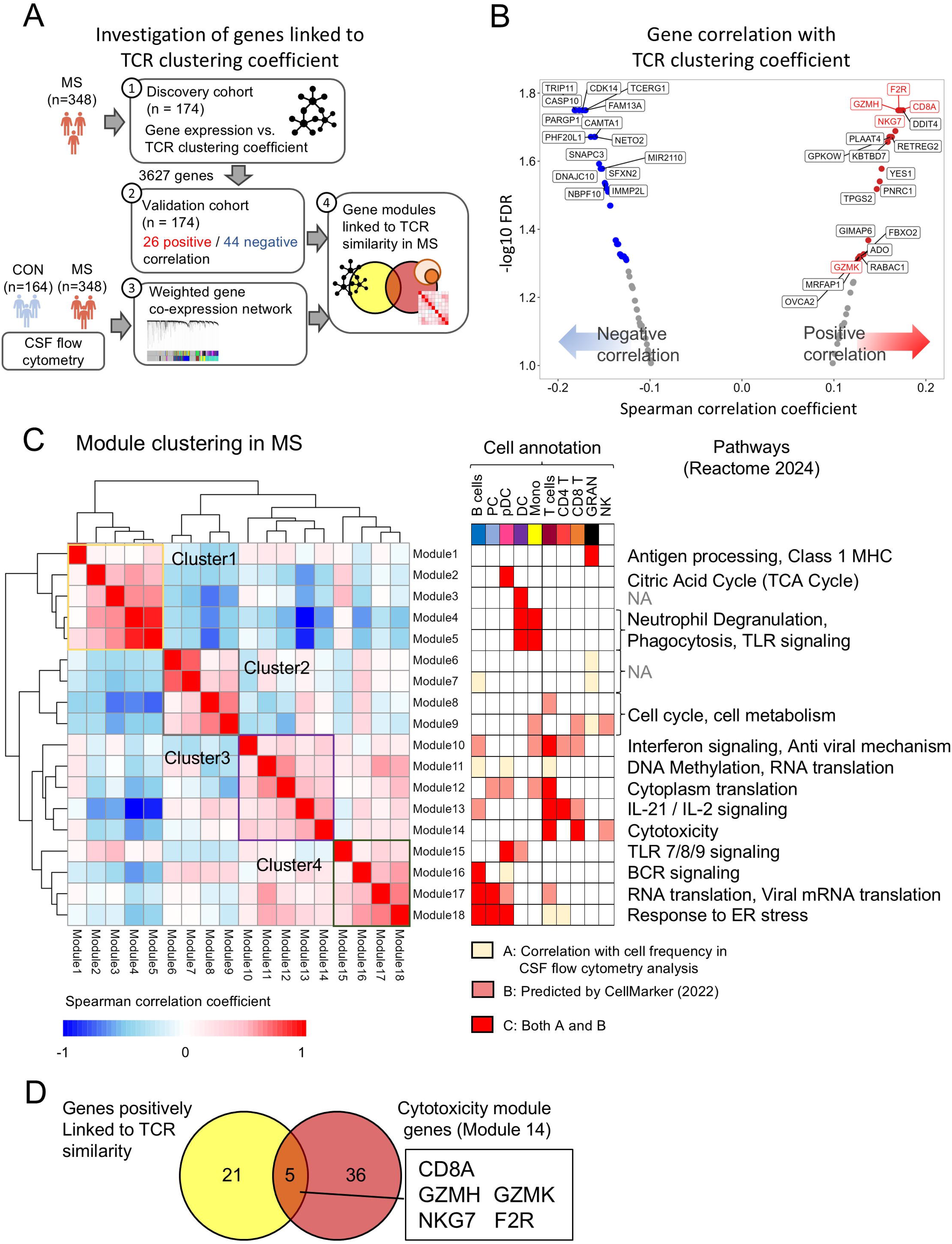
TCR similarity and Bulk RNA seq co-expression network analysis. A. The CDR3 library was generated from cells of pwMS and split into a discovery and validation cohort. Gene expression was compared with TCR clustering coefficient. In the validation cohort, 70 genes showed positive or negative correlations with clustering coefficient. WGCNA (weighted gene co-expression network analysis) was analyzed and control data were used to separate out age and gender effects by regression analysis. The CSF cell proportion in flow cytometry data were compared with WGCNA module score to annotate CSF cell type to each module. B. Volcano plot showed 26 genes with positive correlation with TCR clustering coefficient (red) and 44 genes with negative correlation (blue) in the validation cohort. C. WGCNA generated 18 modules which were clustered by using module eigengene score in MS. Pathway of each module were detected by using Reactome and CSF cell annotation was done as follows: 1. Comparison with cell proportion based on flow cytometric data (yellow) and 2. CellMarker for genes in module (pink). 3. Cell annotation was shown with red if it met both 1 and 2 strategies. D. *CD8A, NKG7, GZMH, GZMK*, and *F2R* were shared between cytotoxicity module (module 14) in WGCNA and genes showing positive correlation with TCR clustering coefficient.

Using weighted gene co-expression network analysis (WGCNA) to reduce dimensionality, we transformed bulk RNA-seq data into 18 modules (Extended Data Figure 3A) which we annotated to immune cell subtypes (see Methods). Correlation identified four module clusters (Figure 3C): Cluster 1 included modules 1-5 annotated to innate immune cells: granulocytes (module 1), plasmacytoid dendritic cells (pDC; module 2), myeloid DC and monocytes (modules 3-5). Cluster 1 negatively correlated with cluster 2 (modules 6-9), which included metabolic and cell cycle pathways without dominant cellular annotations (i.e., shared by cell types). The remaining 2 clusters were annotated to lymphocytes. Cluster 3 comprised modules 10-14 preferentially expressed in T cells, without clear distinction between CD4+ and CD8+ T cells apart from cytotoxicity module 14, which annotated to CD8+ T cells and NK cells. Finally, cluster 4 contained modules 15-18, annotated to B cells (modules 16-18), PC (modules 17-18) and pDC (modules 15&18).

Five genes (*CD8A, NKG7, GZMH, GZMK*, and *F2R*) from cytotoxicity module 14 overlapped with TCR similarity-enriched genes (Figure 3D) and prompted us to search if they are co-expressed in specific CD8+ T cells. To do this, we used single cell RNAseq (scRNA-seq).

### ScRNA-seq of CSF cells identifies GZMH+/GZMK+ CD8+ T cells as unique anti-viral effectors that are clonally expanded and differentiated in MS CSF and enriched for TCRs recognizing EBV epitopes

We aggregated CSF scRNA-seq data from six publications^24,36–40^ (34 MS, 12 controls) with internal CITE-seq cohort (13 MS, 5 controls; Extended Data Table 2) that permitted simultaneous single cell analysis of surface proteins. TCRs from 47,217 T cells were generated from 5’ libraries from 20 pwMS (Figure 4A). Reference annotation^41^ (Extended Data Figure 4A) confirmed increased clonality of CD8+ compared to CD4+ T cells (Extended Data Figure 4B).

**Figure 4:**
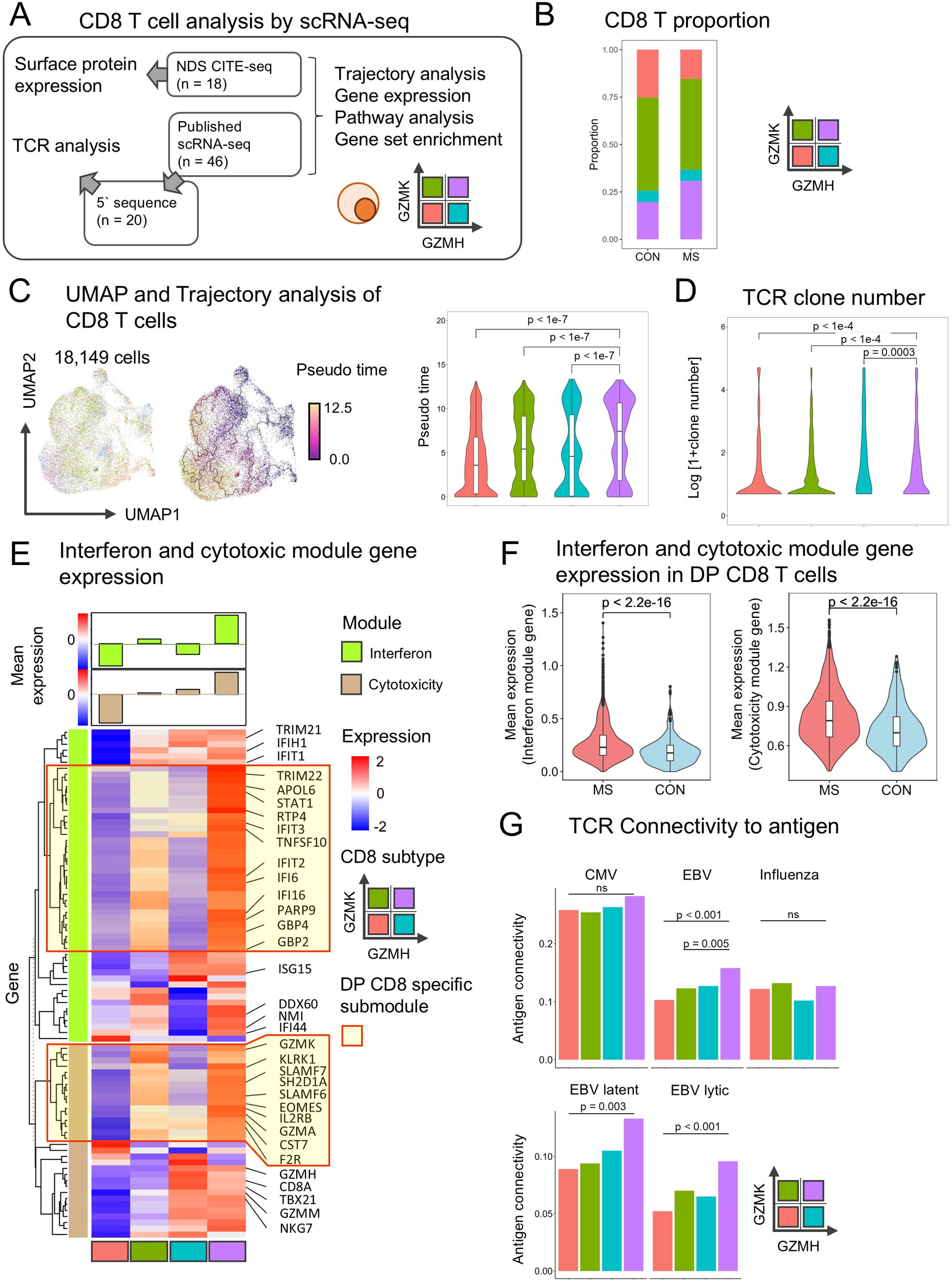
single cell RNA sequencing analysis of CD8 T cells. A. Published single cell (sc) RNA-seq (n=46) and CITE-seq (n=18) were merged for CSF cell analysis. Among published scRNA-seq, 5’ sequence data (MS, n=20) were used for generating TCR libraries. CD8 T cells were split into four groups based on GZMK and GZMH expression. B. Proportion of CD8 T cell subgroup was compared between MS and control. C. Dimension plot of CD8 T cell subgroup and plot with trajectory analysis were shown (left). GZMH/GZMK double positive (DP) CD8 T cells showed highest pseudo time among CD8 T cells (right). D. TCR clone number was analyzed in MS. DP CD8 T cells exhibited highest TCR clonality among CD8 T cells. E. Interferon and cytotoxicity module gene expression was analyzed and average gene expression was assessed for each module. DP CD8 T cells showed highest interferon and cytotoxicity module gene expression. F. Average gene expression of interferon (left)/cytotoxicity (right) module was assessed in DP CD8 T cells and compared between MS and control. G. TCR connectivity to antigen (virus) was analyzed in MS TCR library. DP CD8 T cells showed highest connectivity to EBV (upper) and both EBV latent / lytic proteins (bottom) among CD8 T cells.

Dividing 18,149 CD8+ T cells into 4 categories based on the *GZMH* and *GZMK* expression (Extended Data Figure 4C) showed an increased proportion of double positive (DP) and a decreased proportion of double negative (DN) CD8+ T cells in pwMS (Figure 4B).

Based on a pseudotime trajectory analysis, DP T cells are most- and DN T cells are least differentiated (Figure 4C). DP T cells are also most clonally expanded in pwMS (Figure 4D).

DP cells had the highest (and DN cell had the lowest) expression of Cytotoxicity and Interferon signaling modules (Figure 4E), even though not all module genes were enriched in DP cells.

Unsupervised clustering identified DP CD8+ T cells enriched submodules that are highlighted by yellow shading in Figure 4E. DP cells derived from pwMS had significantly higher Cytotoxicity and Interferon signaling transcripts than DP cells from controls (Figure 4F).

Thus, DP CD8+ T cells are not only more expanded, but also more differentiated anti-viral effectors in pwMS compared to controls, prompting the question of whether they recognize EBV antigens. While TCRs recognizing CMV and Influenza epitopes were equally distributed among all CD8+ T cells, EBV TCRs were significantly enriched in DP cells (Figure 4G). This was true for both latent and lytic EBV epitopes.

We conclude that GZMH/GZMK co-expression identifies intrathecal CD8+ T cells expanded and differentiated in pwMS, with upregulated antiviral immune mechanisms and enriched for TCRs recognizing EBV epitopes.

Our CITE-seq cohort identified a unique cell surface phenotype of DP T cells (Figure 5A). Consistent with their in-vivo activation, DP cells had highest surface levels of activation markers HLA-DR, ICOS and PD-1 and lowest levels of CD127 (IL7RA), downmodulated in recently activated T cells. However, they also lacked CD25, (IL2RA) which is generally upregulated on activated lymphocytes (Figure 5A). Intriguingly, such atypical CD127^-^/CD25^-^ CD8+ T cells are expanded in EBV-mediated infectious mononucleosis^42^.

**Figure 5:**
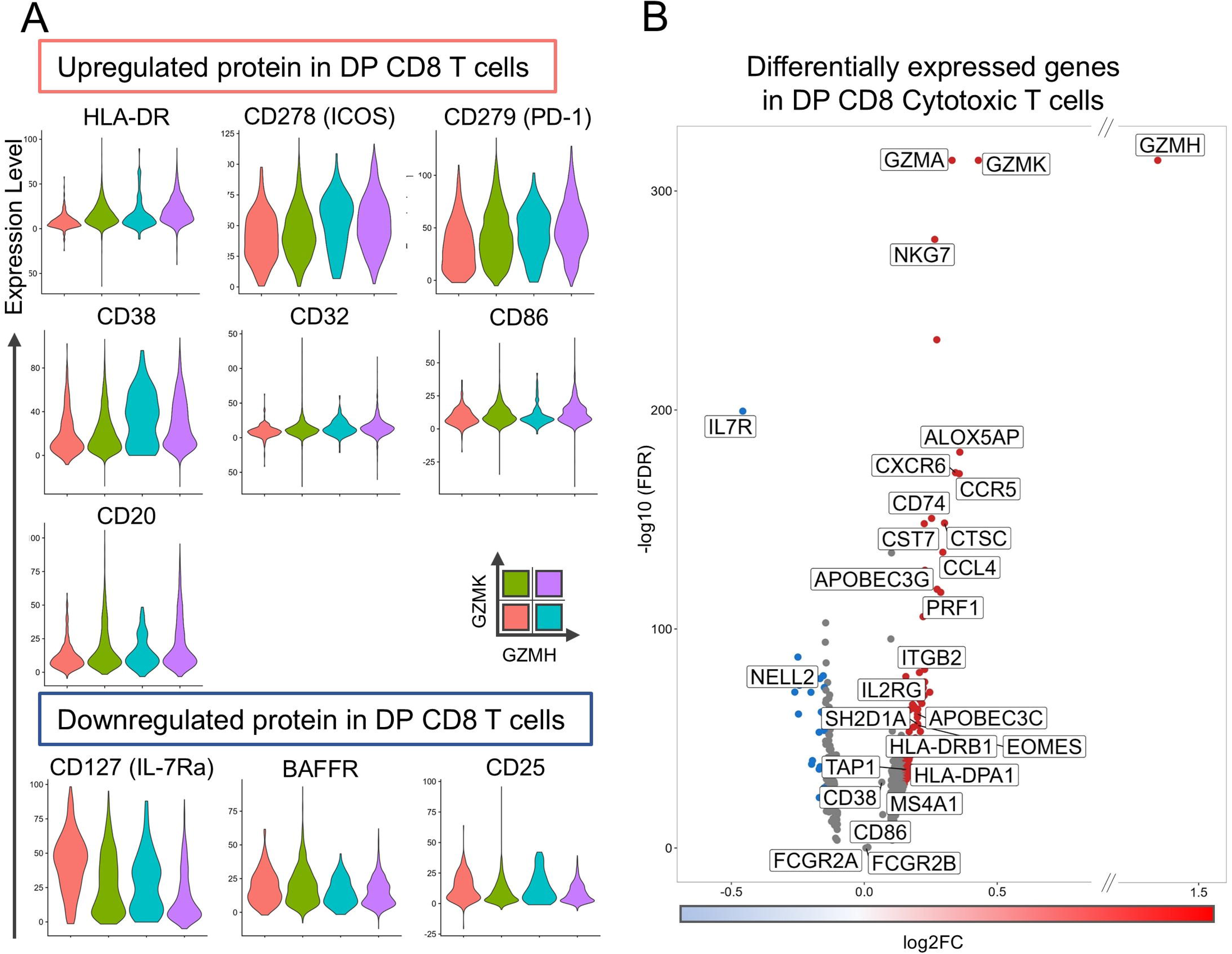
Differential protein and gene expression of DP CD8 T cells. A. Surface protein was analyzed by using CITE-seq data. HLA-DR, ICOS, PD-1, CD38, CD32, CD86 were upregulated in DP CD8 T cells (upper). CD127, BAFF receptor and CD25 were downregulated in DP CD8 T cells (bottom). B. Gene expression was compared between DP CD8 T and other CD8 T cells. Red and blue dots showed significantly upregulated or downregulated genes respectively.

DP T cell membranes also contained molecules not typically expressed in CD8+ T cells that were not transcriptionally upregulated in DP cells (Figure 5B). These included CD32 (*FCGR2A/FCGR2B*), CD86, CD38 and CD20 (*MS4A1*). As these molecules are either uniquely (CD20) or preferentially (CD38, FCGR2B) expressed in B cells and PC, they might have been acquired by DP T cells via trogocytosis^43^, during immune synapse formation.

In addition to high expression of *NKG7, GZMK* and *GZMH*, DP cells also express high levels of *GZMA*, *PRF1* and *CTSC*, affirming their high cytotoxicity. High expression of chemokine markers *CXCR6*, *CCR5*, and *CCL4* (Figure 5B) is consistent with a tissue-resident phenotype of DP cells.

Upregulation of CD74 (CLIP peptide), TAP1, HLA-DRB1, HLA-DPA1 indicates that DP cells can process and present antigens (Extended Data Figure 4D). Finally, DP CD8+ T cells also upregulated genes with an important role in anti-EBV immunity, such as *SH2D1A* (SAP protein)^44^ and *APOBEC3G & −C*^45^.

Thus, the CITEseq results confirm the unique phenotype of MS-expanded DP CD8+ T cells as intrathecal anti-EBV effectors.

### CD8+ T cells expand upon co-culture with autologous EBV-infected CSF B cell lines (EBV-BCL), kill the B cells and the degranulated CD8 T cells acquire a phenotype overlapping with CSF DP CD8+ T cells

The phenotype of the DP cells suggests that they are expanded in pwMS in response to intrathecal EBV infection and that they limit EBV spread by killing B cells that reactivate virus.

To assess this hypothesis, we infected CSF B cells with exogenous EBV, transforming them into immortalized B cell lines (EBV-BCL) as described^46^. From 2 pwMS we managed to immortalize EBV-BCL that differed in EBV reactivation rates measured as the viral load in culture supernatants (Extended Data Figure 5A). This allowed testing T cell responses to autologous EBV-BCL that had either a low (LR-EBV-BCL) or high reactivation rate (HR-EBV-BCL).

First, we analyzed peripheral blood mononuclear cells (PBMC) co-cultured for 7 days with or without cell-free EBV or with autologous LR-EBV-BCL and HR-EBV-BCL (Figure 6A; gating strategy in Extended Data Figure 5B). CD8+ T cells proliferated only when PBMC were co-cultured with EBV-LCL, but not in response to cell-free EBV (Figure 6B). Co-culture with HR-EBV-BCL induced much stronger CD8+ T cell proliferation compared to LR-EBV-LCL co-culture (Figure 6C).

**Figure 6:**
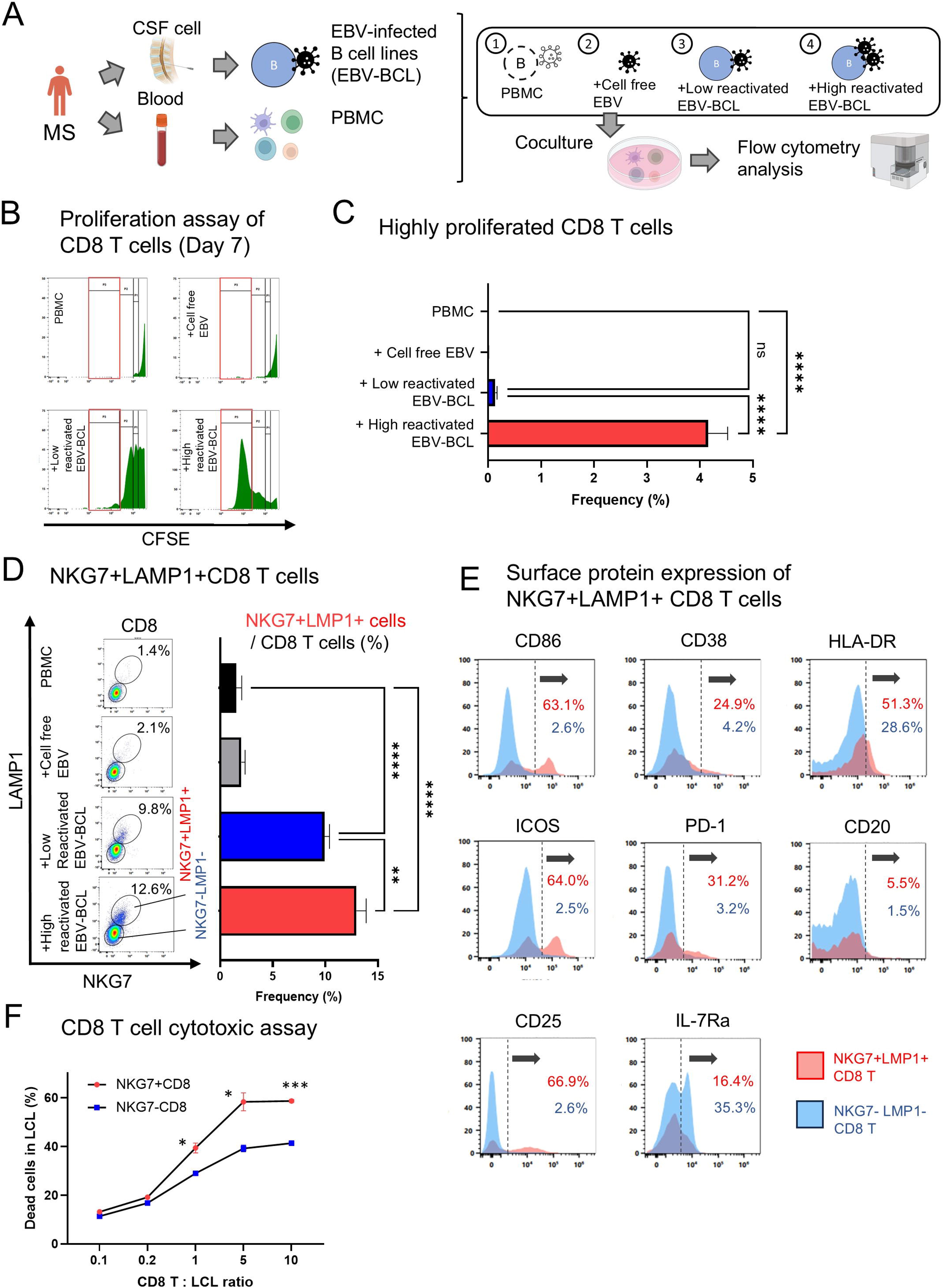
PBMC cocultured with cell free EBV and CSF-EBV cell line. A. PBMC was cocultured with cell free EBV, or low reactivated or high reactivated EBV infected B cell lines (EBV-BCL) from the same patient with MS. After seven days of coculture, PBMC were analyzed by using flow cytometry. B. Proliferation of CD8 T cells in day7 was assessed by using carboxyfluorescein succinimidyl ester (CFSE). Highly proliferated CD8 T cells were gated by red outline. C. PBMC cocultured with EBV-BCL, especially high reactivated EBV-BCL, exhibited high proliferation of CD8 T cells. D. Left plot showed NKG7 / LAMP1 expression among CD8 T cell gate. The frequency of NKG7+/LAMP1+ cells in CD8 T cells increased in PBMC cocultured with EBV-BCL, especially high reactivated EBV-BCL (right). E. Surface protein expression was shown with red (NKG7+LAMP1+ CD8 T cells) or blue color (NKG7-LAMP1-CD8 T cells). F. PBMC was cocultured with high reactivated EBV-BCL for 2 days, and NKG7+ or NKG7-CD8 T cells were sorted. Sorted CD8 T cells were cocultured with EBV-BCL for 6 hours and viability of EBV-BCL were measured by propidium iodide. p-value; *: p < 0.05; **: p < 0.01; ***: p < 0.001, ****: p < 0.0001.

Surface levels of LAMP1 identified CD8+ T cells that degranulated in response to EBV-LCL (Figure 6D, left). LAMP1 surface levels increased on CD8+ T cells co-cultured with EBV-LCL (but not cell free EBV) and LAMP1 colocalized on the cell membrane with NKG7, a transcriptional marker of DP CD8+ T cells. Degranulating, LAMP1+/NKG7+ CD8+ T cells were significantly expanded upon PBMC co-culture with HR-EBV-LCL compared to LR-EBV-LCL (Figure 6D, right).

We asked whether LAMP1+/NKG7+ CD8+ T cells have an overlapping phenotype with DP cells intrathecally expanded in pwMS (Figure 6E). Compared to remaining CD8+ T cells (blue), the LAMP1+/NKG7+ T cells (red) had significantly higher surface levels of CD86, CD38, HLA-DR, ICOS, PD-1 and CD20, and lower CD127 (Extended Data Figure 5C). Thus, co-culture with autologous EBV-BCL undergoing viral reactivation expands CD8+ T cells that degranulate and assume the cell surface phenotype of CSF DP cells. The only exception was CD25, which we found increased on LAMP1+/NKG7+ CD8+ T cells but decreased on CSF DP cells.

However, when we isolated NKG7+ CD8+ T cells after 7-day co-culture with EBV-BCL to formally assess their cytotoxicity, they expressed low levels of GZMK and GZMH (Extended Data Figure 5D) and did not kill EBV-BCL. Hypothesizing that this low cytotoxicity was due to cytotoxic granule depletion after serial killing, we repeated cytotoxicity experiments with NKG7+ versus NKG7-CD8+ T cells isolated after 48-hour co-culture with EBV-BCL. This short co-culture was necessary to enrich rapidly proliferating, degranulating T cells without completely depleting cytotoxic granules, as NKG7+ T cells are practically absent in resting PBMC. Using increasing effector:target (E:T) ratios we observed greater killing of HR-EBV-BCL by NKG7+ as compared to NKG7-CD8 T cells (Figure 6F).

We conclude that CD8+ T cells with a cell surface phenotype analogous to DP CD8+ T cells recognize and kill autologous EBV-infected B cells undergoing lytic reactivation.

### Co-expression modules correlate with development of new/contrast-enhancing lesions (CEL) and are affected by MS disease-modifying treatments (DMTs)

An additional way to investigate the hypothesis that intrathecally expanded DP cells may be beneficial to reduce the severity of MS is by correlating phenotypes of these cells with MS clinical/imaging outcomes and investigating effects of MS disease-modifying treatments (DMTs).

While co-expression modules did not correlate with any clinical outcome (not shown), lymphocyte-related modules correlated with CELs (Figure 7A; note that because we did not obtain post-contrast MRI with every CSF, correlations with CELs identified on brain MRI were limited to 215 pwMS. Therefore, we used validated CSF proteomic-CEL prediction^47^ to maximize use of pwMS with matched CSF-clinical data). The correlations with CELs were stronger in untreated pwMS, likely due to an inhibitory effect of MS DMTs on CEL formation. However, while B cell-related modules and some T cell-related modules still correlated with CELs in treated pwMS, the cytotoxicity module that partially reflected DP CD8+ T cells ceased to correlate with CELs in treated pwMS.

**Figure 7:**
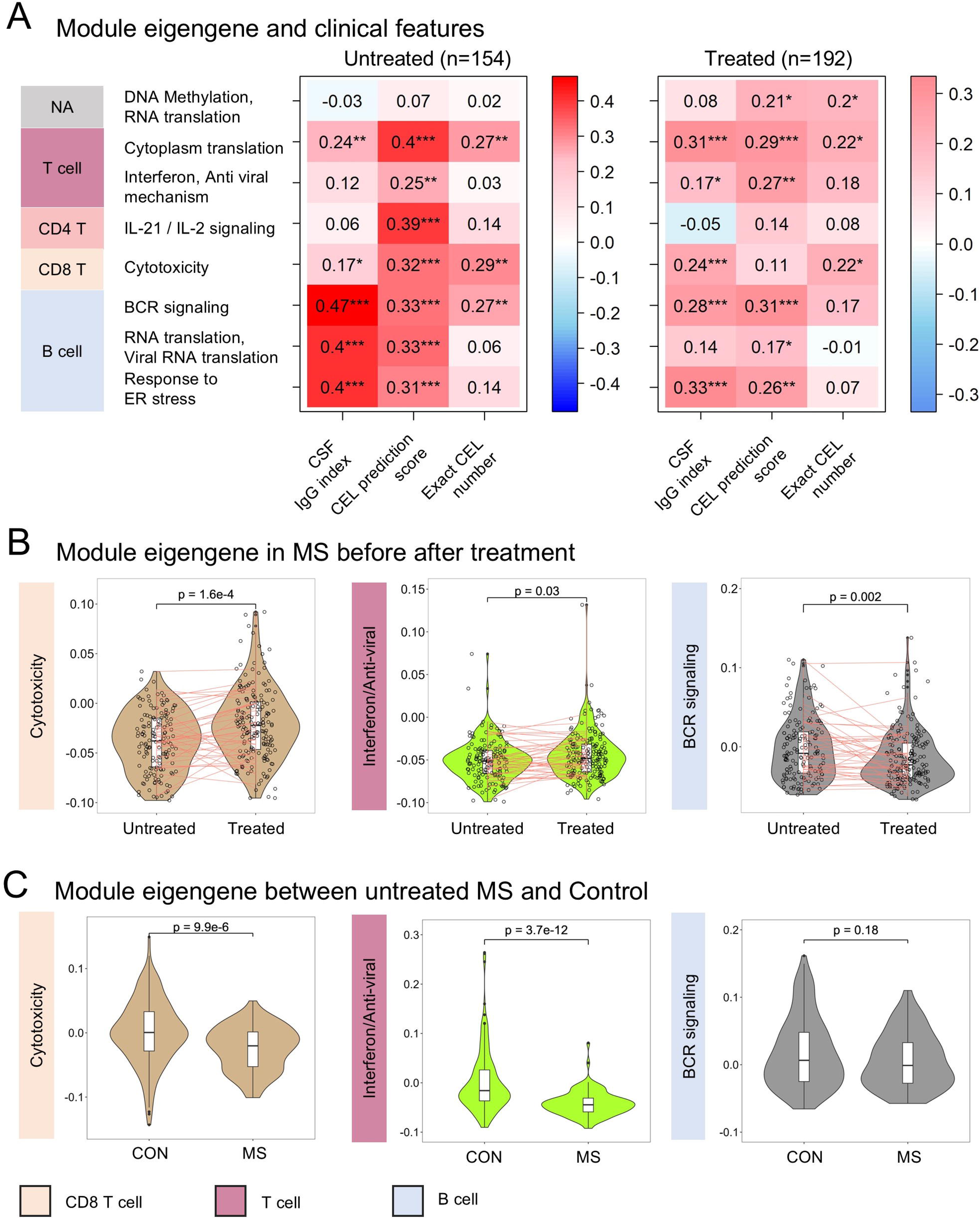
WGCNA module score associated with contrast-enhancing MS lesions (CEL) and are affected by MS disease-modifying treatments (MS-DMTs) A. Bulk RNA-seq WGCNA module score was compared with CSF IgG index, MS contrast-enhancing lesions (CEL) prediction score, and exact contrast-enhancing MS lesion number in MRI. CEL score was generated by CSF protein prediction model for CEL. Spearman’s rank correlation coefficient was shown with p-value; *: p < 0.05; **: p < 0.01; ***: p < 0.001. B. WGCNA module score was compared between MS with and without disease-modifying treatments (DMTs). Red line indicated samples from same MS patients. The Mann-Whitney U test was used for comparisons between two groups. C. WGCNA module score was compared between untreated MS and control. Cytotoxicity/Interferon modules were lower in untreated MS compared with control.

As expected, the IgG index that measures intrathecal IgG production also positively correlated with B cell/PC modules in untreated pwMS, but RNA translation/viral RNA translation module did not correlate in treated pwMS. The CD8+ cytotoxicity module (and T cell-related cytoplasmic translation module) also correlated with IgG index. This B cell/T cell correlation between co-expression modules was already noted in Figure 3C and Extended Data Figure 6A.

As MS DMTs inhibit formation of MS lesions (measured by CELs) we asked whether DMTs also affect co-expression modules. Compared to untreated pwMS, treated patients had significantly lower BCR signaling module and higher cytotoxicity and interferon signaling modules (Figure 7B, Extended Data Figure 7A). The matched before-after treatment CSF samples visualized by connecting red lines demonstrated congruent effects. The CD8 cytotoxicity and interferon modules were significantly lower in untreated MS compared to controls (Figure 7C), supporting the hypothesis of an impaired T cell response to viral infections in MS.

While correlation is not causation, a hypothesis that best integrates our observations is that intrathecal B cell lytic EBV reactivation initiates MS lesion formation, inducing activation/expansion of anti-EBV T cells, which limit intrathecal EBV spread. This hypothesis yields two predictions: (1) anti-EBV T cell responses expand proportionally to CELs, as CELs reflect the amount of lytic EBV reactivation, (2) MS outcomes depend on the balance between anti-EBV T cells and EBV-reactivating B cells akin to an E:T ratio that determines if the former population contains the latter, and (3) DP CD8+ T cells have a protective role in MS disability progression.

To test these predictions we used linear regression models between B cell RNA translation/viral RNA translation modules and T cell interferon signaling (Figure 8A,B) and between BCR signaling and cytotoxicity modules (Figure 8C,D) as these showed the strongest B-T cell correlations (Extended Data Figure 6A).

**Figure 8:**
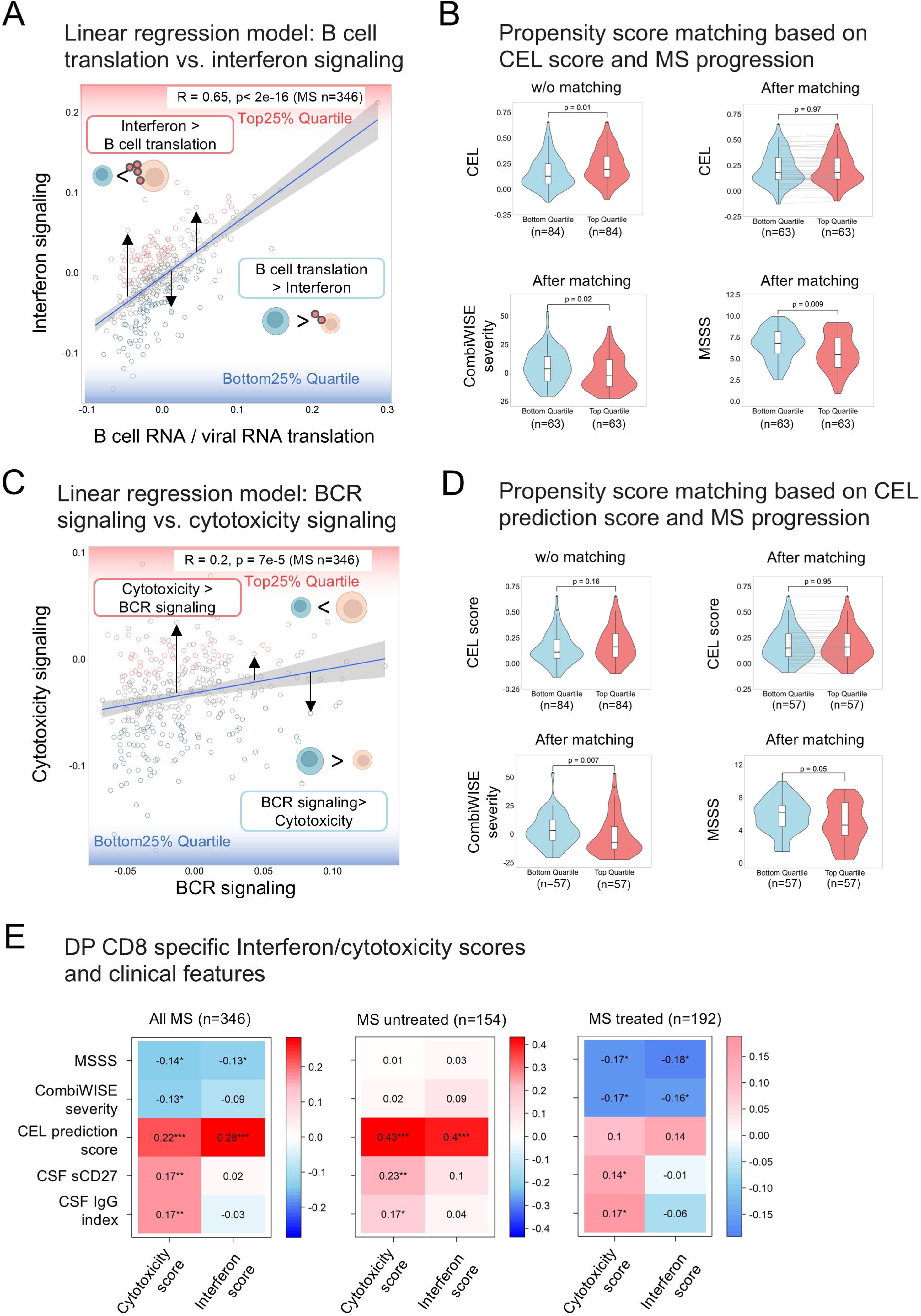
pwMS with Cytotoxicity and IFN-signaling dominance to B cell module exhibited lower MS severity after separating out CEL effects by regression analysis and propensity score matching. A. Linear regression model indicated the association between B cell RNA/viral RNA translation and interferon signaling. Black arrow indicated the residuals of the linear regression model. Red color dots showed samples with top 25% quartile of residuals and blue dots belonging to bottom 25% quartile of residuals. B. CEL score was compared between top/bottom 25 % quartile groups (B cell transcription vs. interferon signaling model) before and after propensity score matching based on CEL score (upper). CombiWISE severity and MSSS were compared between top/bottom 25 % quartile groups after propensity score matching. C. Linear regression model indicated the association between B cell receptor (BCR) signaling module and cytotoxicity signaling module scores. Red color dots showed samples with top 25% quartile of residuals and blue dots belonging to bottom 25% quartile of residuals. D. CEL score was compared between top/bottom 25 % quartile groups (BCR signaling vs. cytotoxicity signaling model) before and after propensity score matching by using CEL score (upper). combiWISE severity and MSSS were compared between top/bottom 25 % quartile groups after propensity score matching. E. DP CD8 T cell specific interferon/cytotoxicity scores were calculated based on sum of submodule of each interferon or cytotoxicity gene expression. Each score was compared with CSF IgG index, soluble CD27, CEL score and MS severity (CombiWISE severity, MSSS) in all MS, MS with treated/untreated. Spearman’s rank correlation coefficient was shown with p-value; *: p < 0.05; **: p < 0.01; ***: p < 0.001.

Dividing pwMS into quartiles based on regression model residuals identified patients with proportionally higher (Figure 8B,D; top quartile, red) versus lower (bottom quartile, blue) T cell modules when adjusting for B cell modules. Consistent with prediction #1, pwMS in the top quartile had significantly higher CELs than pwMS in the bottom quartile.

To eliminate the confounding role CELs exert on T cell modules, we used propensity score matching to select subgroups of subjects from top and bottom quartiles with comparable CELs (Figure 8B,D “after matching” panels). We then asked whether pwMS with proportionally higher T cell responses (top quartile) differ from pwMS with proportionally lower T cell responses in rates of disability accumulation measured by MS severity outcomes. The most common MS severity outcome, the Multiple Sclerosis Severity Score (MSSS)^48^ relates disability measured by Expanded Disability Status Scale (EDSS, 0-10 range, ordinal) to MS duration. More granular CombiWISE severity relates disability measured by the Combinatorial Weight-Adjusted Disability Scale^49^ (CombiWISE; 0-100 range, continuous and correlating with EDSS: Rho = 0.98, p<0.0001^49^) to subject’s age. Consistent with prediction #2, pwMS with proportionally higher interferon signaling (Figure 8B lower panels) and cytotoxicity modules (Figure 8D lower panels) had significantly lower MS severity.

Finally, to use all pwMS CSF data but focusing on DP CD8+ T cells, we computed DP cell enriched submodules of cytotoxicity and interferon signaling modules (Figure 4E, yellow highlighted) and investigated their relationship to MS outcomes (Figure 8E). DP cell-enriched submodules correlated positively with CELs and negatively with MS severity outcomes in pwMS (Figure 8E, left panel). Their positive correlation with CELs was observed solely in untreated pwMS (Figure 8E, middle panel) while negative correlations with MS severity outcomes were observed in treated pwMS (Figure 8E, right panel). MS-DMTs significantly increased DP CD8 specific Cytotoxicity/Interferon submodules in MS (Extended Data Figure 7B). This supports prediction #3, that DP CD8+ T cells limit rate of MS disability accumulation.

## Discussion

This decade-long study of the CSF cell transcriptome in a large cohort of pwMS validated key results from previous smaller studies and provided crucial novel observations extending our understanding of MS immunopathogenesis.

Compared to controls, pwMS have chronic intrathecal activation of adaptive immune responses, evidenced by significantly higher BCR and TCR clonality. Nevertheless, both BCR and TCR CSF responses in pwMS are polyclonal with a high overlap between pwMS and controls, implying that MS-specific T and B cell responses represent small subpopulations of the CSF repertoire.

Combining TCR repertoire analysis with TCR connectivity and WGCNA modules annotated to CSF immune cells in the large bulk RNAseq cohort helped us to identify 5 co-expressed cytotoxic genes, raising a possibility that they are co-expressed in a single cell type. We confirmed this by scRNAseq datasets identifying these cells as GZMH+/GZMK+ CD8+ T cells that are activated, clonally expanded, and differentiated in pwMS compared to controls. Their phenotype is consistent with unique anti-viral cytotoxic effectors, and they are enriched for TCRs annotated to recognize EBV epitopes. CMV and influenza annotations showed that this TCR enrichment is specific for EBV, comprising both latent and lytic epitopes, with the latter being more prominent in pwMS. We also show that PBMC co-culture with autologous CSF EBV-BCL activates and expands CD8+ T cells that assume a surface molecule phenotype overlapping with DP CD8+ T cells expanded in MS CSF and that these cells can kill EBV-BCL proportionally to the extent of their EBV reactivation.

Although indirectly, these results imply ongoing intrathecal EBV infection in pwMS, including lytic reactivation. The only alternative explanation for persistent clonal expansion and activation of anti-EBV cytotoxic CD8+ T cells in MS would be their cross-reactivity to CNS antigens.

However, that possibility is excluded by negative correlations of DP cells enriched cytotoxicity and interferon signaling co-expression submodules with MS severity outcomes. In other words, if these cells cross-recognized CNS auto-antigens, their intrathecal expansion should lead to faster, not slower disability accumulation. Our results support a beneficial role for anti-EBV DP CD8+ T cells in MS, inferring a pathogenic role of persistent EBV infection in MS progression.

We discussed already in the introduction practical considerations that may prevent consistent detection of EBV in MS CNS by current pathology techniques. In view of these limitations we believe that negative studies do not rule out EBV-mediated immunopathogenesis in MS progression.

Because B cell -depleting treatments abrogate CEL formation with comparable or higher efficacy than all other immunomodulatory MS drugs, B cells may be primary driver of MS lesion formation. EBV establishes latent infection in B cells, but the virus circulates between B cells and epithelial cells, causing episodic lytic infection with virus shedding in the latter^50^. Linking our interim results with published observations that EBV can infect also (brain) endothelial cells^51^ led us to hypothesize that a subpopulation of latently infected B cells in pwMS are opening the blood brain barrier (BBB) by mediating EBV lytic infection of brain endothelial cells and that CSF T cells expanded in response to this intrathecal EBV infection may limit MS-associated CNS injury due to their cytotoxicity and interferon-related effector mechanisms. While this hypothesis cannot be studied in living pwMS directly, we derived 3 testable inferences.

Analyzing relationships between relevant B cell and T cell co-expression modules and use of propensity score matching to eliminate confounding effect of CELs helped us to show that all 3 inferences are consistent with our hypothesis.

While our study does not rule out a contributing role of autoimmunity (perhaps Ab or CD4+ T cells mediated) in MS progression, it strongly implies a role for primary EBV-mediated CNS immunopathogenesis, by mechanisms that need to be elucidated in future studies.

During preparation of this manuscript, we became aware of the results in preprint form from independent investigators^52^, studying non-overlapping pwMS using very different techniques. They identified an analogous population of CD8+ T cells expanded in pwMS CSF and show that some of these cells recognize EBV antigens, but not autoantigens. These combined results provide strong support that EBV not only mediates MS onset but persists in the MS CNS where periodic lytic virus reactivation contributes to CNS damage and accumulation of disability.

## Online methods

### Study design

All subjects participating in Neuroimmunological Diseases Section of NIAID/NIH natural history protocol “Comprehensive Multimodal Analysis of Neuroimmunological Diseases of the Central Nervous System” (Clinicaltrials.gov identifier NCT00794352) between 2012 and 2023 were included. All subjects signed informed consent and the protocol was approved by NIH IRB. Up to 6/2022, CSF cells (1×10e4) were analyzed fresh by 12 color flow cytometry to identify 14 subpopulations of immune cells as described^53^. Bulk RNA from the remaining CSF cells (if ≥1×10e4) was sequenced in 3 batches. After 6/2022 flow cytometry with bulk RNAseq was replaced by cellular indexing of transcriptome and epitopes (CITESeq) in pwMS with ≥ 1×10e4 CSF cells, providing simultaneously singe cell RNAseq (scRNAseq) and quantitation of surface proteins by sequencing. All sequencing data were generated on coded samples by investigators blinded to the clinical/ imaging data.

### Sample collection

For bulk RNA-seq of CSF cells, 495 MS patients (145 with RRMS, 131 with SPMS, and 219 with PPMS), 18 CIS patients, and 276 controls (170 with OIND, 58 with NIND, and 48 with HD) were prospectively recruited from November 2012 to December 2018. The inclusion criteria for HD cohort were ages 18–75 years, lack of neurological abnormality or systemic disease or brain MRI. For single cell RNA-seq of CSF cells, 13 MS subjects (5 with RR-MS, 2with SP-MS, 6 with PP-MS) and 5 controls (1 with NIND and 4 HD) were prospectively recruited from August 2022 to March 2023.

### CSF Sample processing

CSF was collected on ice and processed according to a written standard operating procedure by investigators blinded to diagnoses, clinical, and imaging outcomes. Aliquots were assigned alphanumeric identifiers and centrifuged for 10 minutes at 300g at 4°C within 30 minutes of collection. The pelleted CSF cells were processed by flow cytometry or processed for bulk or single cell RNA sequencing. For bulk RNA sequencing, CSF cells were stored with RNA protection at −80 degree.

### Flowcytometric analysis

Fresh CSF cells collected from ∼20cc of CSF were resuspended in 200ul of X-VIVO TM 15541(Lonza, REF: 04-418Q) on ice. A minimum of 2,000 cells were stained with a 12-color antibody panel, analyzed by BD Bioscience LSR II flow cytometer as described (citation). The results have been prospectively entered into the research database, quality-controlled and locked from further changes. The database automatically calculated proportions of each cell type based on total cell number.

### Bulk RNA sequencing

Prior to RNA extraction and NGS library preparation samples were balanced and randomized based on diagnostic condition, gender, and CSF collection date in a 96-well plate format. All sample processing was performed in 96-well format by one individual. CSF cells were centrifuged at 5,000 x *g* for 15 minutes in a swinging-bucket Tomy Micro Twin High Speed centrifuge. Pelleted cells were combined with 700uL RLT buffer and genomic DNA and RNA were extracted following AllPrep DNA/RNA 96-well centrifugation protocol with the additional on-column DNAse 1 treatment. RNA quality was assessed using Agilent’s 2100 Bioanalyzer and the RNA Pico 6000 kit (Agilent Technologies, Santa Clara, CA). RNA quantity was low in most the samples, which necessitated CSF RNA quantitation by QRTPCR. We used AgPath-ID One-Step RT-PCR Buffer and Enzyme Mix (Life Technologies, Carlsbad, CA). The reactions were carried out in 20 µL reactions using human Beta-2 microglobulin (B2M) forward primer (5’-GCCGTGTGAACCATGTGACTT-3’), reverse primer (5’-AAATGCGGCATCTTCAAACCT-3’), and fluorescent probe (5’-TGATGCTGCTTACATGTCTCGATCCCACTT-3’; Biosearch Technologies, Novato, CA). The QPCR reactions were carried out at 50 °C for 10 minutes, 95 °C for 10 minutes, 55 cycles of 95 °C for 15 seconds and 60 °C for 45 seconds. Data were analyzed using ABI 7900HT version 2.4 sequence detection system software (Life Technologies, Carlsbad, CA). RNA quantity per sample was determined by absolute quantitation method (Thermofisher Scientific, Waltham, MA).

The libraries were prepared using SMARTer Stranded Total RNA-seq V3 Pico Input Mammalian protocol. The libraries were pooled and sequenced on one NovaSeq 6000 S2 flowcell using 2×101 cycles paired-end sequencing. The Illumina RTA v3.4.4 was used to process raw data files, the Illumina bcl2fastq2.17 was used to demultiplex and convert binary basecalls and quality scores to fastq format. The sequencing reads were trimmed adapters and low-quality bases using Cutadapt (v 1.18). The trimmed reads were mapped to human reference genome (hg38) and the annotated transcripts (GENCODE v30) using STAR aligner (v2.5.1) with two-pass alignment option. RSEM (version 1.2.11) was used for gene and transcript quantification based on GENCODE annotation file.

### Single cell RNA sequencing and raw data processing

The pelleted CSF cells were processed by flow cytometry or processed for single cell RNA sequencing. Fresh CSF cells were collected as described in sample processing. Cells were labelled with 79 Totalseq B antibodies (Biolegend, USA) shown in the table (Extended Data Table 3) and washed with PBS with 1% Bovine Serum Albumin (Thermo Fisher Scientific, USA).

Labeled cells were fixed by using fixed RNA feature barcode kit (10x Genomics, USA) and stored at −80 degree. Cellular indexing of transcriptomes and epitopes by sequencing (CITE-seq) method was used to analyze transcriptome and surface protein information based on single-cell readout. After probe hybridization, gene expression library and surface protein library were constructed by using Chromium Next GEM Single cell 3’ LT Reagent kits v3.1 (Dual index) (10X Genomics, USA) following the protocol (CG000400). Single-cell fixed RNA libraries and single-cell feature barcode libraries were sequenced on a NextSeq 2000 P3 28 cycle + 90 cycle asymmetrical run. Demultiplexing yielded over 27 million reads per sample when allowing for 1 mismatch per index adapter.

### RNA sequencing data processing

In single cell RNA-seq data analysis, raw data were counted, and QC was checked by Cellranger ver.7.0 (10X Genomics). Published data of scRNA-seq of CSF cells from MS and control (GSE133028, GSE163005, GSE138266, PRJNA866296, GSE172003, GSE138266)^24^ ^36^ ^37–40^was recruited. Doublet cells were excluded by using DoubletFinder ver.2.0^54^ and protein library from CITE-seq was denoised by using dsb package^55^. The cells with high mitochondrial gene (>10%) or high unique molecular identifier (UMIs) (>8,000) were excluded. Filtered out Read count data were as merged by using Canonical correlation analysis (Seurat V4, reciprocal PCA method)^41^.

The merged dataset was clustered by Seurat, K-nearest neighbor (KNN) graph and UMAP. Non-biased clustering was done by using weighted nearest neighbor analysis and annotated by using human PBMC data^41^. Trajectory analysis was done by Monocle3^56^. Pathway analysis was done using IPA®(QIAGEN, USA).

### BCR and TCR analyses and CDR3 network visualization

For single cell and bulk RNA sequencing, TCR and BCR library was generated by using TRUST4^57^. BCR libraries were generated from 789 bulk RNA-seq data of CSF cells using TRUST4. 258 were excluded due to complete lack of BCR data and five samples were filtered out as outliers. This left 348 MS samples, 164 control samples, and 14 clinically isolated syndrome (CIS) samples for further analysis. TCR repertoire data were generated for these 526 samples. TCR annotation to antigen was analyzed by using published TCR dataset (VDJdb: https://vdjdb.cdr3.net/, McPAS-TCR: http://friedmanlab.weizmann.ac.il/McPAS-TCR/, and TBAdb: https://db.cngb.org/pird/).

We categorized TCR specificity predictions from these databases into four categories: 1. Foreign microbes, 2. Cancer, 3. Autoimmune disease, and 4. Other antigens^58^.

We used Levenshtein distance to assess the distance between two Complementarity-determining regions 3 (CDR3) amino acids and defined similarity as *exp (-lev.distance (CDR3[i], CDR3[j])).* We randomly selected 1,500 CDR3s shared by ≥ 2 subjects in each library of MS specific, control specific, and shared between MS and control to generate a TCR network. Using these 4,500 CDR3s for nodes and their similarities for connection weight, we constructed a TCR CDR3 network. We quantified network similarity by calculating the clustering coefficient (Cu):

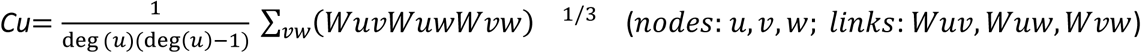

When plotting the association between TCR clone number and average clustering coefficient, we observed that CSF samples with low TCR numbers (<50 TCRs; Extended Data Figure 2C) had high variance of clustering coefficient, implying noise. Thus, we compared clustering coefficient and Edge density only for CSF samples with TCR clone number ≥50. Network X (ver. 3.4.2)^59^ in Python3.5 was used to generate CDR3 network and Cytoscape (ver. 3.10.3)^60^ was used to visualize the CDR3 network. Edge density was calculated to assess tightness of connection in each cluster as follows:

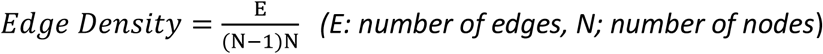

### Bulk RNA sequencing network analysis

For network analysis, weighted gene co-expression network analysis (WGCNA)^61^ was used for generating. We removed all genes with counts < 15 in more than 75% of samples. Outliner of samples were excluded based on dendrogram (Extended Data Figure 3B). Module eigengene (ME) is calculated as the first principal component of the expression matrix of each module. To account for gender and age effects on MEs, we compared each module ME with age and gender using control samples (Extended Data Figure 3D). Age effects were filtered from Module10 and 14, and gender effects were regressed out from Module5 (Extended Data Figure 3D) and applied to MS samples. Spearman correlation coefficients were calculated for MEs, and ME modules were clustered based on Distance = 1 – Spearman correlation coefficient. Each module was annotated to specific CSF cell types by comparing with flow cytometric data (R≥0.2, p<0.05)using CellMarker (2024) (FDR<0.05)^62^. Genes included in each module was analyzed by using Reactome (2024)^63^.

### EBV cell line and PBMC culture

PBMCs were collected from peripheral blood by using Vacutainer Mononuclear Cell Preparation Tube (BD Bioscience, Cat. 362761) and stored in X-Vivo (Lonza, Cat. 02-060Q) containing 20% FBS (Gibco, Cat. 16000044) and 10% DMSO at liquid nitrogen tank. Lymphoblastoid cell lines (LCLs) were generated by seeding CSF cells on Irradiated (6,000 Rd) CD40+ NIH 3T3 with EBV containing supernatants from B95-8 cells, 2.5ug/mL CpG ODN 2006 (InvivoGen, Cat. tlrl-2006), interferon-γ (IFN-γ), and 0.5 µg/ml Cyclosporine A (Sigma Aldrich, Cat. C1832-5mg) as described in our previous report^46^. LCLs were cultured in 25mL T flask (4×105 cells/mL) with RPMI 1640 Medium (Cat. 11875093), 10% Fetal bovine serum (Cat. 26140079), Amphotericin-B (1μg/mL, Cat. 15290018) (all from Thermo Fisher), and penicillin (100 U/mL, streptomycin (100μg/mL) (Sigma Aldrich, Cat. P4333-20ML). DNA of LCL was extracted by using DNA kit (QIAGEN, USA) and EBV DNA was quantified by using BALF5 (Primer1: 5’-CCCTGTTTATCCGATGGAATG-3’, Primer2: 5’-CGGAAGCCCTCTGGACTTC-3’) with ZenQuench system (Integrated DNA Technologies, USA) and RT-PCR (BioRad, USA). X50-7 cell line was used for standard following previous reports (J Virol. 2009 Sep 9;83(22):11857–11861). PBMC (10^6^ cells) was cocultured with LCL (10^4^ cells) from same patient or cell free EBV (B95.8) in 2mL X-Vivo for 7 days. Cultured cells were stained with antibodies (Extended Data Table 4) and analyzed by Cytek Aurora and SpectroFlo (Cytek Bioscience, USA). In proliferation assay, PBMC was labelled by using CellTrace™ CFSE Cell Proliferation Kit (Invitrogen™, Cat. C34570) following the protocol and CFSE signal of CD8+ cells were analyzed. In cytotoxicity assay, PBMC were cocultured with LCL for 48 hours following the above methods. After that, CD8 T cells were sorted by using CD8+ T Cell Isolation Kit (130-096-495; Miltenyi Biotec, USA) and split into NKG7+ and NKG7-CD8 T cells with NKG7 Rabbit mAb (clone:E6S2A, Cell signaling technology, USA) and Anti-Rabbit IgG MicroBeads (130-048-602; Miltenyi Biotec, USA). LCL (2×10^5^, 10^5^, 2×10^4^, 4×10^3^, 2×10^3^ cells) labelled with CD19 were cocultured with sorted NKG7+ or NKG7-CD8 T cells for 6 hours. Cell viability of LCL was assessed by using Propidium iodide (#4830, R&D systems, USA).

### Clinical outcomes and CEL prediction Score

Combinatorial Weight Adjusted Disability Scale (CombiWISE) showing strong correlation with Expanded Disability Status Scale^49^, and MS Disease Severity Scale (MSSS)^48^ as MS progression scores. We used propensity score matching (Matching ver.4.1)^64^ to filter out the effect of CEL on MS progression score, resulting in top - bottom quartile paired samples. CEL score was predicted through the model generated by using CSF somamers (Novartis V3-5K, SomaLogic Inc, Boulder, CO, USA) and CEL number as an outcome^65^.

### Statistical analyses

Mann-Whitney U test was used for comparisons between two groups, while One-way ANOVA and Tukey-Kramer test were applied for comparisons among three or more groups. The p-values were FDR (false discovery rate)-adjusted for multiple comparisons across gene expression. We used Spearman’s rank correlation to compare two values. Chi-squared test was used to compare reference-annotated TCR frequency between pwMS and control. All statistical analyses were performed using RStudio software (ver. 4.1.3)^47^.

Extended Data Figure legends

Extended Data Figure1

A. The frequency of IGHV (IGHV4.31 and IGHV4.59) was compared between MS patients with and without HLA risk allele (HLA.DRB1.15:01 or 15:03). B. Linear regression model depicting the relationship between total TCR read counts and total TCR clone numbers. C. CDR3 library was annotated to specific antigen by using three CDR3 cohorts (VDJdb, McPAS-TCR, and TBAdb). Antigens were classified into four groups: 1. Foreign microbes, 2. Cancer, 3. Autoimmune disease, and 4. Other antigens. Set size and intersection size indicated the number of TCRs.

Extended Data Figure2

A. The antigen annotation: The frequency of HLA-A02 restricted TCRs specific for CMV, EBV, and influenza were compared between MS and control. TCR from pwMS showed higher frequency of TCRs annotation to EBV (left). The frequency of HLA-A02 restricted TCRs annotation to lytic and latent EBV protein were assessed and compared between MS and control (right). Lytic protein was predicted to annotated higher in MS than control. B. Clustering coefficient of TCRs with specificity for CMV, EBV, and influenza were analyzed and compared between MS and control. Clustering coefficient of TCRs recognizing EBV latent or lytic tended to elevated in MS. The Mann-Whitney U test was used for comparisons between two groups. C. Linear regression model showed the relation between TCR clone number and average clustering coefficient (left) and Distribution of average clustering coefficient (right, red dot and error bar showed mean and standard deviation, respectively). The linear model was used to define cut-off level of TCR clone number. R indicated clustering coefficient.

Extended Data Figure3

A. Scale-free topology network generated 18 modules. B. Outliner of samples were excluded based on dendrogram. C. Power level of scale-free topology network was decided by using scale topology model fit. D. Interferon/anti-viral module score and cytotoxicity module score correlated with age in control. Phagocytosis/TLR module score showed the correlation with Female in control. Each linear regression model was used to regress out age or gender effect in module score in MS. R indicated clustering coefficient.

Extended Data Figure4

A. Dimension plot showed 158,283 CSF cells annotated to PBMC reference. B. CD8 T cells indicated higher clonality compared with control in MS CSF. C. Ridge plot showed *GZMH* and *GZMK* expression. Cells with *GZMH* ≥ 1 or *GZMK* ≥ 1 were defined as *GZMH*+ cells or *GZMK*+ cells, respectively. D. Subclusters of interferon and cytotoxicity module genes indicated genes highly expressed in *GZMH*+*GZMK*+ Double positive (DP) CD8 T cells. D. Pathway analysis was analyzed based on differential gene expression between GZMH/GZMK DP and other CD8 T cells.

Extended Data Figure5

A. EBV DNA copy number of EBV-B cell line (BCL) generated form pwMS was analyzed by using qPCR (n=2). B. Gating strategy of PBMC with and without cell free EBV or EBV-BCL. B. Surface protein expression of CD8 T cells. Red bar showed NKG7+LAMP1+ CD8 T and blue one was NKG7-LAMP1-CD8 T cells. D. Intracellular GZMH and GZMK protein expression was assessed in CD8 T cells after 7 day coculture with and without cell free EBV or EBV-BCL. P-value was adjusted by FDR; **: p < 0.01; ***: p < 0.001.

Extended Data Figure6

A. Correlation matrix of WGCNA module score was generated by using all MS samples (top, n=346), MS with treatment (middle, n=154), and without treatment (bottom, n=192). Spearman’s rank correlation coefficient was shown with p-value; *: p < 0.05; **: p < 0.01; ***: p < 0.001.

Extended Data Figure7

A. WGCNA module score was compared between untreated and treated MS patients. Red line indicated samples from same subject. B. Submodules specific for GZMH/GZMK double positive (DP) CD8 T cells were defined based on clustering of gene expression in single cell RNA-sequencing data. Cytotoxicity / Interferon submodule scores were compared between untreated and treated MS. Red line indicated samples from same subject.

## Data Availability

All data produced in the present study are available upon reasonable request to the authors

## Acknowledgments

This research was funded by the Division of Intramural Research, NIAID/NIH. We thank Kennichi Dowdell for providing cell-free EBV.

