## Supplementary figures and images for "Intrathecally expanded GZMK+/GZMH+ CD8 T cells targeting EBV antigens may reduce severity of Multiple Sclerosis"

### ExFig1.tiff

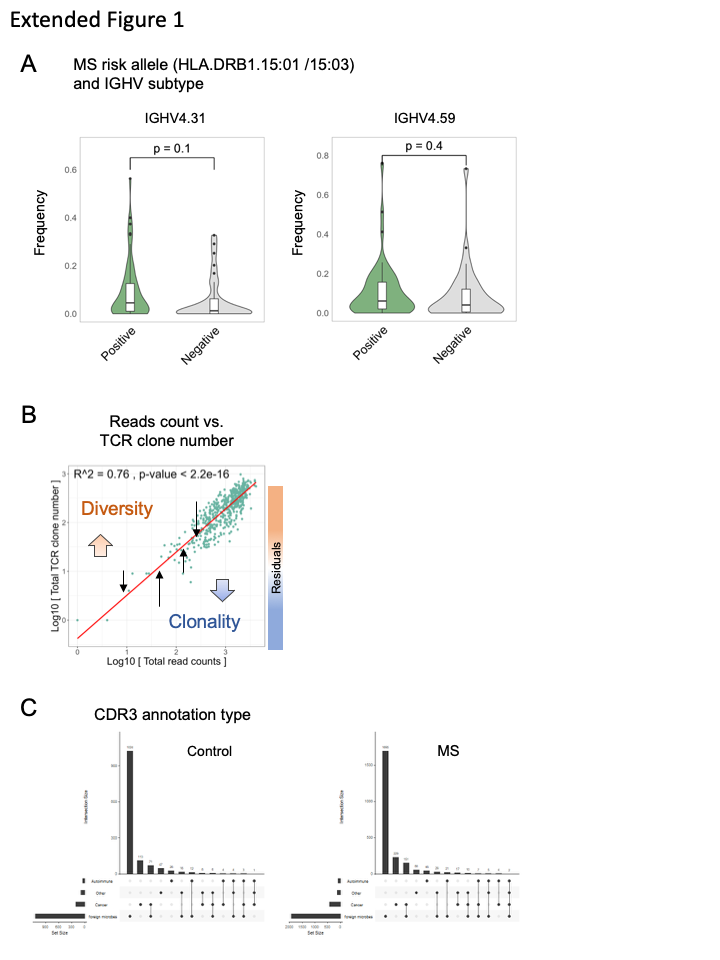

### ExFig2.tiff

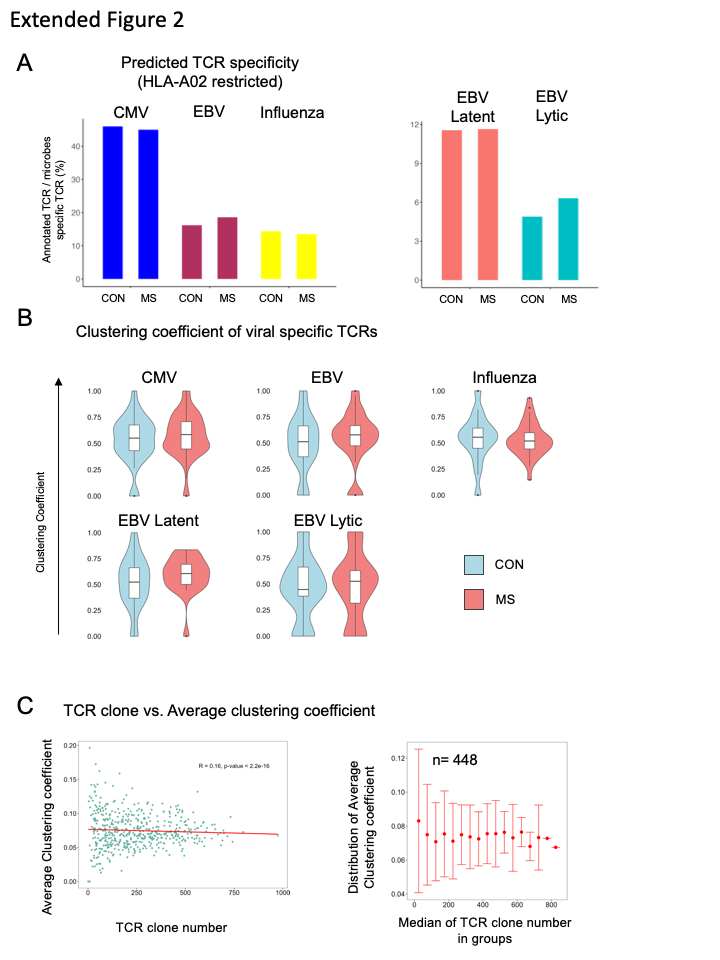

### ExFig3.tiff

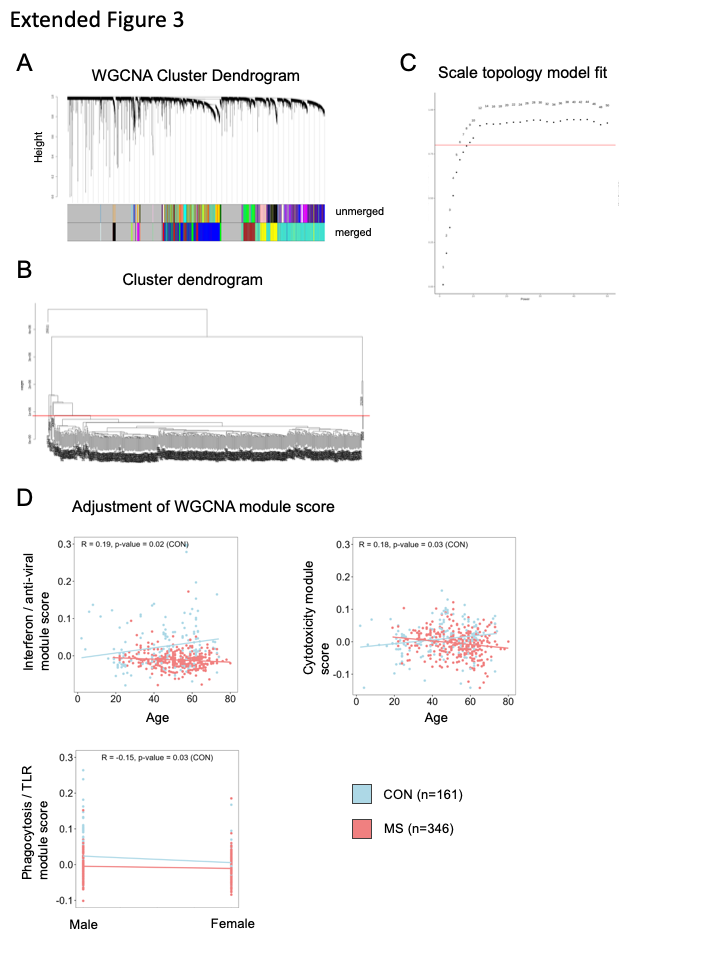

### ExFig4.tiff

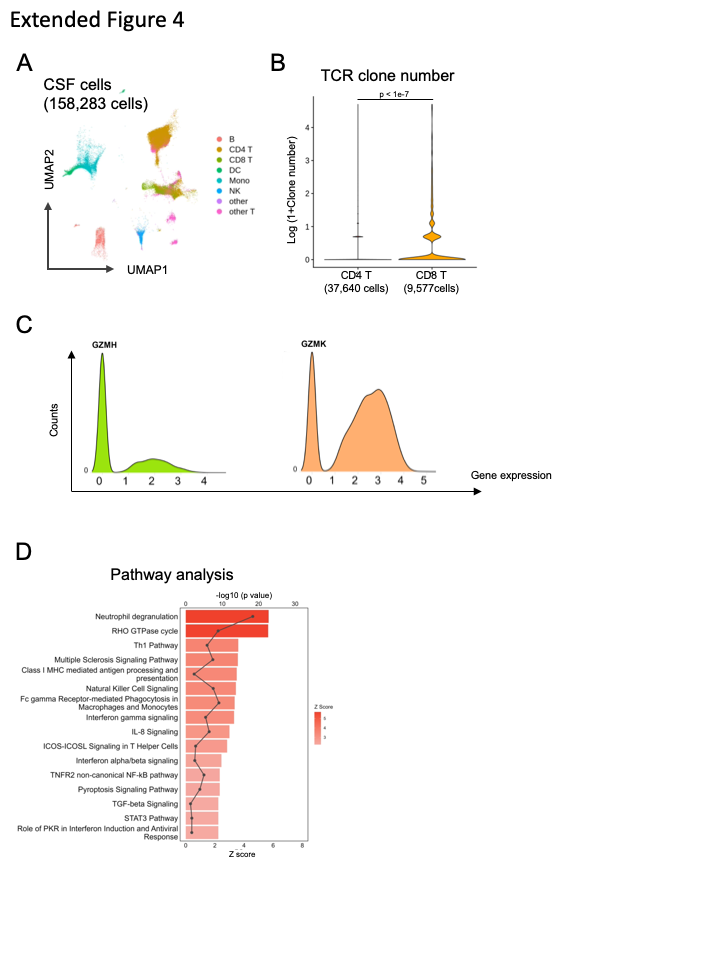

### ExFig5.tiff

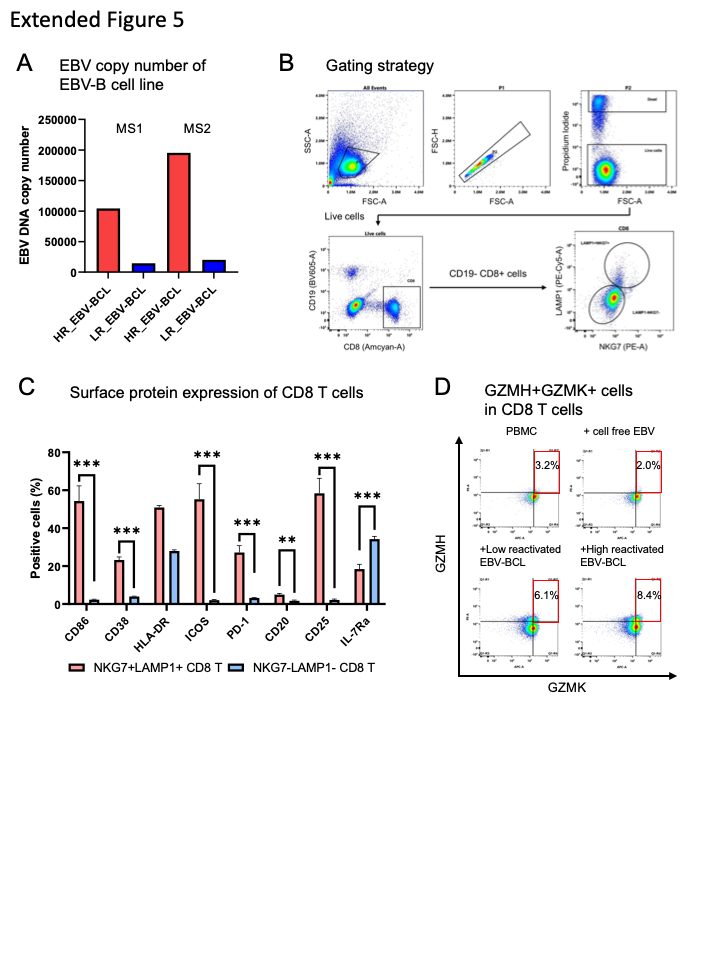

### ExFig6.tiff

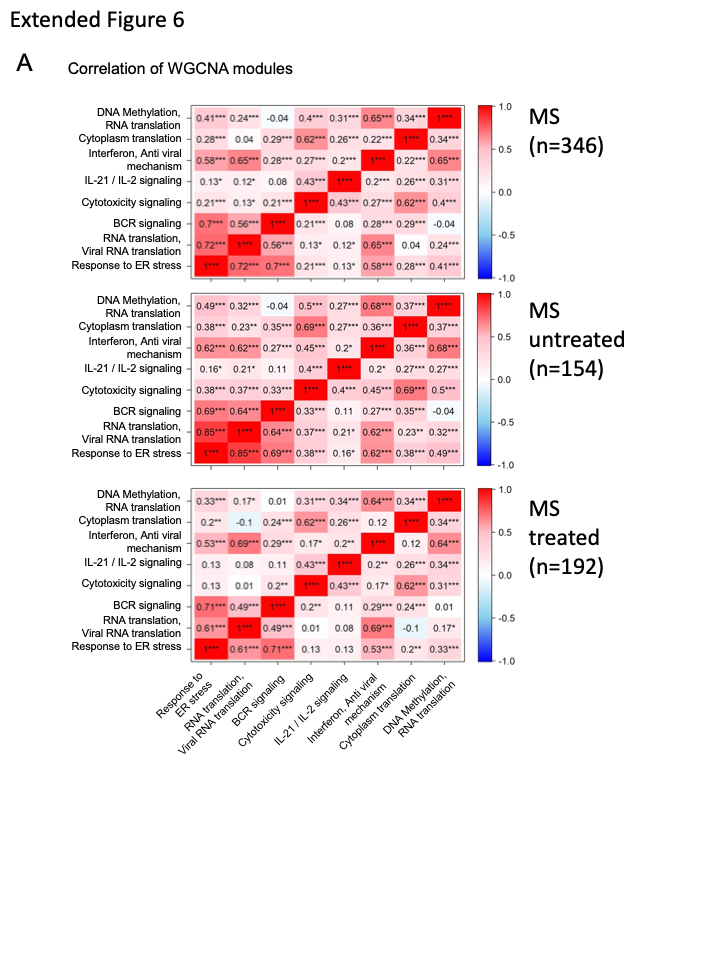

### ExFig7.tiff

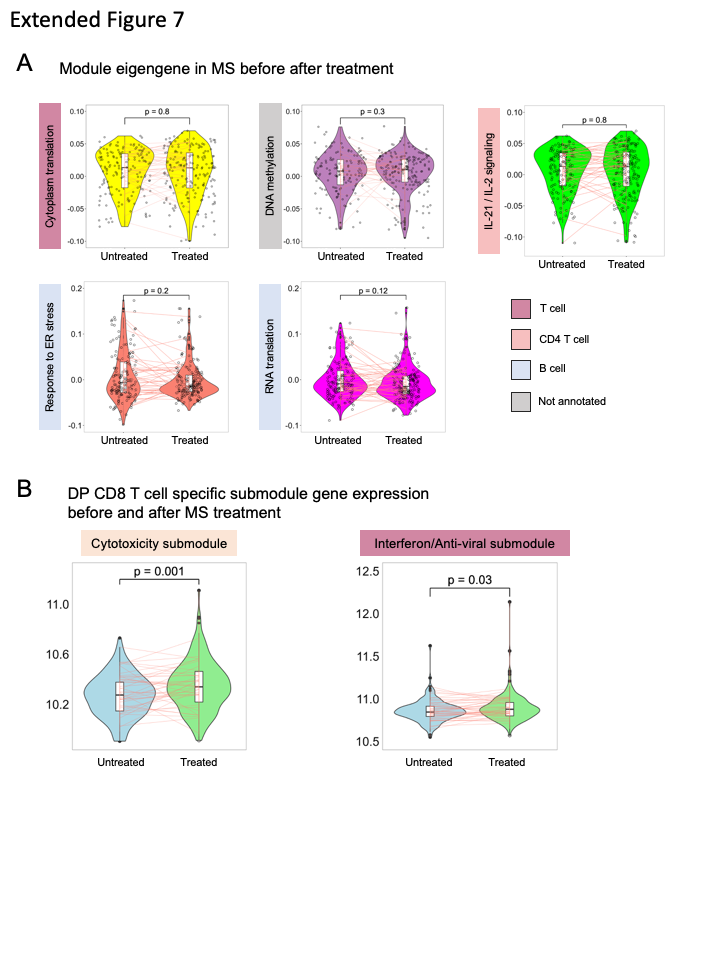
